# Racial and ethnic disparities in COVID-19 vaccinations in the United States during the booster rollout

**DOI:** 10.1101/2021.12.12.21267663

**Authors:** Jeremy Samuel Faust, Benjamin Renton, Utibe R. Essien, Céline R. Gounder, Zhenqiu Lin, Harlan M. Krumholz

## Abstract

**Background:** We sought to quantify whether there were statistically significant disparities along race and ethnicity lines during the early rollout of Covid-19 vaccine booster doses in the United States. We also studied whether such disparities replicated or widened disparities that had already been observed during the initial series rollout as of 2 months earlier (Janssen) or 6 months earlier (Pfizer-BioNTech or Moderna), which comprised the booster-eligible population.

**Methods:** This cross-sectional study of US adults (ages ≥18 years) used public data from US Centers for Disease Control and Prevention. The observed shares of vaccine doses for each race and ethnicity were compared to the expected shares, predicted based upon the compositions of the booster-eligible and initial series-eligible populations.

**Results:** As of November 16, 2021, 123.5 million US adults were eligible for a booster dose of either the Pfizer-BioNTech, Moderna, or Janssen vaccines. Of these, 21.7 million had received a booster dose, among whom race and ethnicity information was available for 18.8 million booster recipients.

A statistically significant higher share of Non-Hispanic White and Non-Hispanic Multiple/Other race individuals had received a booster vaccination than projected based on the composition of the booster-eligible population. A statistically significant lower share of Hispanic, Non-Hispanic American Indian/Alaskan Native, Non-Hispanic Asian, Non-Hispanic Black, and Non-Hispanic Native Hawaiian/Other Pacific Islander individuals had received a booster vaccination than expected based on the booster-eligible population. A secondary analysis of the booster-eligible population found that some of these disparities had already occurred at the time of the initial series. However, the booster campaign widened all of those disparities and added new disparities for Non-Hispanic American Indian/Alaskan Native and Non-Hispanic Native Hawaiian/Other Pacific Islander individuals.

**Conclusion:** Disparities in Covid-19 vaccine administration on race and ethnicity lines occurred during the initial series rollout in the US. However, these disparities were not merely replicated but widened by the early booster rollout.

## Introduction

US Centers for Disease Control and Prevention (CDC) data reveal marked disparities in Coronavirus disease-2019 (COVID-19) vaccine administration by race and ethnicity.^1,2^ We sought to quantify these disparities and determine whether they widened during the booster rollout.

## Methods

Using CDC publicly available (with the exception of Texas) vaccination and population data,^3,4^ race and ethnicity data were categorized into 7 groups: Hispanic/Latino and Non-Hispanic (NH) American Indian/Alaskan Native (AI/AN), Asian, Black, Native Hawaiian/other Pacific Islander (NHOPI), White, and Multiple/Other race. For each group, initial vaccination administration (by May 16, 2021 for the Pfizer-BioNTech and Moderna vaccines, and September 16, 2021 for the Janssen vaccine) was compared by race and ethnicity with each group’s share of the vaccine-eligible adult population (US adults ages ≥18). Then, the share of booster doses administered (by November 16, 2021) to recipients of known race and ethnicity was compared with each group’s share of the booster-eligible population (US adults ages ≥18 who had been vaccinated 6 months earlier if receiving Pfizer-BioNTech or Moderna vaccines, or 2 months earlier if receiving Janssen vaccines). A statistically significant disparity was defined as any observed share of vaccine doses falling below the 95% CI for that group’s projected share, based on the composition of the eligible populations. Analyses were performed in SAS 9.4.

## Results

As of November 16, 2021, there were 123.5 million booster-eligible US adults, among whom 27.1m (21.9%) had received a booster. Race and ethnicity information was available for 18.8m (69.4%) booster recipients. A statistically significant higher share of NH-White and NH-Multiple/Other race individuals had received a booster vaccination than expected based on the composition of the booster-eligible population (Figure, Table). A statistically significant lower share of Hispanic, NH-AI/AN, NH-Asian, NH-Black, and NH-NHOPI individuals had received a booster vaccination than expected based on the booster-eligible population (Figure, Table).

To assess whether disparities in booster administration replicated or widened ones present during the initial series rollout, an analysis of the initial series was conducted. As of November 16, 2021, there were 123.5m booster-eligible US adults (ie. had completed the initial series either 2 months (Janssen) or 6 months prior (Pfizer-BioNTech, Moderna)); race and ethnicity information was available for 85.8m (69.5%). Given the composition of the vaccine-eligible population, a statistically significantly higher share of NH-White, NH-AI/AN, NH-NHOPI, and NH-Multiple/Other individuals had completed the initial series and were booster-eligible than expected (Figure, Table); a statistically significantly lower share of Hispanic, NH-Asian, and NH-Black individuals had completed the initial series and were booster-eligible than expected (Figure Table).

## Discussion

These data indicate that disparities in Covid-19 vaccine administration on race and ethnicity lines during the initial series in the US were widened by the early booster rollout. If the booster rollout had not widened existing disparities, the share of booster doses received by each race or ethnicity would have replicated those observed during the initial series. Instead, the US booster campaign widened disparities among Hispanic, NH-Asian and NH-Black individuals and introduced disparities among NH-AI/AN and NH-NHOPI individuals.

Because the booster-eligible population was comprised of individuals who had received the initial series, these findings cannot simply reflect lower vaccine interest.^5^ Alternative explanations include decreased access to care in communities with higher rates of racial and ethnic minority individuals, a known driver of similar disparities.^6^ For example, mass vaccination sites and other community-based programs likely narrowed what otherwise might have been even greater disparities during the initial rollout. However, many such sites were closed during the booster rollout. Also, vulnerable populations may have had less access to booster-related public messaging and therefore been unaware both of their booster eligibility and any corresponding benefits.

While disparities in the administration of the initial series have narrowed for some groups in recent months, current policies require that either 2 months (Janssen) or 6-months (Pfizer-BioNTech and Moderna) must have elapsed before an individual becomes booster-eligible. This policy stands to reinforce the inequities we have described here.

Study limitations include data lags and incomplete reporting of race and ethnicity data.

## Data Availability

All data produced in the present study are available upon reasonable request to the authors

## Data Statement and Author Contributions

Dr. Faust had full access to all of the data in the study and takes responsibility for the integrity of the data and the accuracy of the data analysis.

*Concept and design:* Faust, Renton, Krumholz

*Acquisition, analysis, or interpretation of data:* Faust, Renton, Essien, Gounder, Lin, Krumholz

*Drafting of the manuscript:* Faust, Essien, Krumholz

*Critical revision of the manuscript for important intellectual content:* Faust, Essien, Gounder, Lin, Krumholz

*Statistical plan and analysis:* Faust, Lin

*Administrative, technical, or material support:* Renton

*Supervision:* Faust, Lin, Krumholz

## Funding Statements

This study had no specific funding.

## Conflicts of Interest

Faust: None.

Renton: None.

Essien: Dr. Essien reported receiving grants from the Department of Veterans Affairs. Gounder: None.

Lin: Dr Lin reported working under contract with the Centers for Medicare & Medicaid Services.

Krumholz: In the past three years, Dr Krumholz received expenses and/or personal fees from UnitedHealth, IBM Watson Health, Element Science, Aetna, Facebook, the Siegfried and Jensen Law Firm, Arnold and Porter Law Firm, Martin/Baughman Law Firm, F-Prime, and the National Center for Cardiovascular Diseases in Beijing. He is an owner of Refactor Health and HugoHealth, and had grants and/or contracts from the Centers for Medicare & Medicaid Services, Medtronic, the U.S. Food and Drug Administration, Johnson & Johnson, and the Shenzhen Center for Health Information.

---

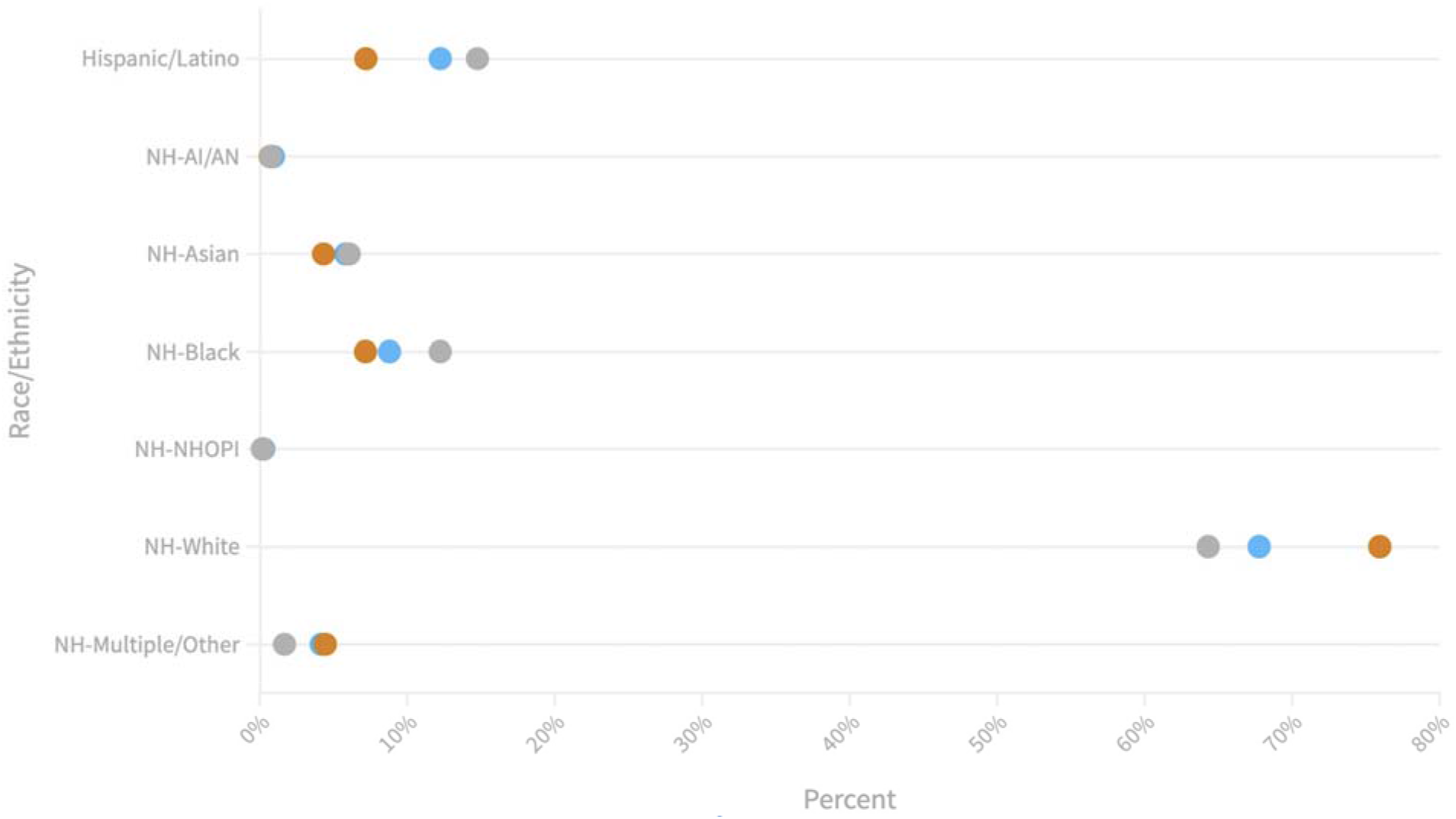
COVID-19 Vaccinations by Race and Ethnicity in the United States, 2021. The gray circles indicate the share of COVID-19 vaccine-eligible US adults; the blue circles indicate the share of COVID-19 vaccine initial series doses received among the eligible US adult population for each race/ethnicity; the orange circles indicate the share of COVID-19 vaccine booster doses received among the booster-eligible US adult population for each race/ethnicity, as of November 16, 2021. In all cases, 95% confidence intervals are synonymous with the visible centers of the corresponding circles, with no overlap between any adjacent or overlapping circles (see Table), and are therefore not shown.

**Table.** COVID-19 Vaccinations and disparities by Race and Ethnicity for the initial series and booster in the United States, 2021

| Race/ethnicity | Adult population | Share of adult population | Vaccinated adults (initial series)* | Share of initial doses received* | Absolute disparity** | Chi-square p-val |
| --- | --- | --- | --- | --- | --- | --- |
| Total (with known race and ethnicity) | 234,736,383 |  | 85,806,925 |  |  | <0.001 |
| Hispanic/Latino | 34,643,869 | 14.76% (95% CI 14.75% - 14.76%) | 10,505,659 | 12.24% (95% CI 12.24% - 12.25%) | -2.52% |  |
| Non-Hispanic AI/AN**** | 1,759,908 | 0.75% (95% CI 0.75% - 0.75%) | 797,976 | 0.93% (95% CI 0.93% - 0.93%) | 0.18% |  |
| Non-Hispanic Asian | 14,255,119 | 6.07% (95% CI 6.07% - 6.08%) | 5,004,914 | 5.83% (95% CI 5.83% - 5.84%) | -0.24% |  |
| Non-Hispanic Black | 28,734,901 | 12.24% (95% CI 12.24% - 12.25%) | 7,547,654 | 8.80% (95% CI 8.79% - 8.80%) | -3.44% |  |
| Non-Hispanic NHOPI***** | 437,957 | 0.19% (95% CI 0.19% - 0.19%) | 231,614 | 0.27% (95% CI 0.27% - 0.27%) | 0.08% |  |
| Non-Hispanic White | 150,959,164 | 64.31% (95% CI 64.30% - 64.32%) | 58,138,817 | 67.76% (95% CI 67.75% - 67.77%) | 3.45% |  |
| Non-Hispanic Multiple/Other | 3,945,465 | 1.68% (95% CI 1.68% - 1.68%) | 3,580,291 | 4.17% (95% CI 4.17% - 4.18%) | 2.49% |  |
| Race/ethnicity | Booster-eligible adult population | Share of booster-eligible population | Vaccinated adults (booster dose)*** | Share of booster doses received*** | Absolute disparity** | Chi-square p-val |
| Total (with known race and ethnicity) | 85,806,925 |  | 18,832,653 |  |  | <0.001 |
| Hispanic/Latino | 10,505,659 | 12.24% (95% CI 12.24% - 12.25%) | 1,355,230 | 7.20% (95% CI 7.18% - 7.21%) | -5.04% |  |
| Non-Hispanic AI/AN**** | 797,976 | 0.93% (95% CI 0.93% - 0.93%) | 134,060 | 0.71% (95% CI 0.71% - 0.72%) | -0.22% |  |
| Non-Hispanic Asian | 5,004,914 | 5.83% (95% CI 5.83% - 5.84%) | 815,801 | 4.33% (95% CI 4.32% - 4.34%) | -1.50% |  |
| Non-Hispanic Black | 7,547,654 | 8.80% (95% CI 8.79% - 8.80%) | 1,351,030 | 7.17% (95% CI 7.16% - 7.19%) | -1.63% |  |
| Non-Hispanic NHOPI***** | 231,614 | 0.27% (95% CI 0.27% - 0.27%) | 36,595 | 0.19% (95% CI 0.19% - 0.20%) | -0.08% |  |
| Non-Hispanic White | 58,138,817 | 67.76% (95% CI 67.75% - 67.77%) | 14,302,149 | 75.94% (95% CI 75.92% - 75.96%) | 8.18% |  |
| Non-Hispanic Multiple/Other | 3,580,291 | 4.17% (95% CI 4.17% - 4.18%) | 837,788 | 4.45% (95% CI 4.44% - 4.46%) | 0.28% |  |

\*As of May 16, 2021 for Pfizer-BioNTech or Moderna, of September 16, 2021 for Janssen
\*\*Negative values indicate a lower share of the corresponding population was vaccinated than expected based on the share of the eligible population; positive values indicate a higher share than expected based on the share of the eligible population.
\*\*\*As of November 16, 2021
\*\*\*\*American Indian/Alaskan Native
\*\*\*\*\*Native Hawaiian/Other Pacific Islander

## Notes

### Funding Statement

This study did not receive any funding

